# Using routine programmatic data to estimate the population-level impacts of HIV self-testing: The example of the ATLAS program in Cote d’Ivoire

**DOI:** 10.1101/2022.02.08.22270670

**Authors:** Arlette Simo Fotso, Cheryl Johnson, Anthony Vautier, Konan Blaise Kouamé, Papa Moussa Diop, Romain Silhol, Mathieu Maheu-Giroux, Marie-Claude Boily, Nicolas Rouveau, Clémence Doumenc-Aïdara, Rachel Baggaley, Eboi Ehui, Joseph Larmarange, the ATLAS team

## Abstract

**Background:** HIV self-testing (HIVST) is recommended by the World Health Organization as an additional HIV testing approach. Since 2019, it has been implemented in Côte d’Ivoire through the ATLAS project, including primary and secondary distribution channels. While the discreet and flexible nature of HIVST makes it appealing for users, it also makes the monitoring and estimation of the population-level programmatic impact of HIVST programs challenging. We used routinely collected data to estimate the effects of ATLAS’ HIVST distribution on access to testing, conventional testing (self-testing excluded), diagnoses, and antiretroviral treatment (ART) initiations in Côte d’Ivoire.

**Methods:** We used the ATLAS project’s programmatic data between the third quarter (Q) of 2019 (Q3 2019) and Q1 2021, in addition to routine HIV testing services program data obtained from the President’s Emergency Plan for AIDS Relief dashboard. We performed ecological time series regression using linear mixed models.

**Findings:** The results are presented for 1000 HIVST kits distributed through ATLAS. They show a negative but nonsignificant effect of the number of ATLAS HIVST on conventional testing uptake (−190 conventional tests [95% CI: −427 to 37, p=0·10]). We estimated that for 1000 additional HIVST distributed through ATLAS, +590 [95% CI: 357 to 821, p<0·001] additional individuals have accessed HIV testing, assuming an 80% HIVST utilization rate (UR) and +390 [95% CI: 161 to 625, p<0·001] assuming a 60% UR. The statistical relationship between the number of HIVST and HIV diagnoses was significant and positive (+8 diagnosis [95% CI: 0 to 15, p=0·044]). No effect was observed on ART initiation (−2 ART initiations [95% CI: −8 to 5, p=0·66]).

**Interpretations:** Social network-based HIVST distribution had a positive impact on access to HIV testing and diagnoses in Cote d’Ivoire. This approach offers a promising way for countries to assess the impact of HIVST programs.

**Funding:** Unitaid 2018-23-ATLAS

**Research in context:** *Evidence before this study:* We searched PubMed between November 9 and 12, 2021, for studies assessing the impact of HIVST on HIV testing, ‘conventional’ testing, HIV diagnoses and ART initiation. We searched published data using the terms “HIV self-testing” and “HIV testing”; “HIV self-testing” and “traditional HIV testing” or “conventional testing”; “HIV self-testing” and “diagnosis” or “positive results”; and “HIV self-testing” and “ART initiation” or “Antiretroviral treatment”. Articles with abstracts were reviewed. No time or language restriction was applied. Most studies were individual-based randomized controlled trials involving data collection and some form of HIVST tracking; no studies were conducted at the population level, none were conducted in western Africa and most focused on subgroups of the population or key populations. While most studies found a positive effect of HIVST on HIV testing, evidence was mixed regarding the effect on conventional testing, diagnoses, and ART initiation.

*Added value of this study:* HIVST can empower individuals by allowing them to choose when, where and whether to test and with whom to share their results and can reach hidden populations who are not accessing existing services. Inherent to HIVST is that there is no automatic tracking of test results and linkages at the individual level. Without systematic and direct feedback to program implementers regarding the use and results of HIVST, it is difficult to estimate the impact of HIVST distribution at the population level. Such estimates are crucial for national AIDS programs. This paper proposed a way to overcome this challenge and used routinely collected programmatic data to indirectly estimate and assess the impacts of HIVST distribution in Côte d’Ivoire.

*Implications of all the available evidence:* Our results showed that HIVST increased the overall HIV testing uptake and diagnoses in Côte d’Ivoire without significantly reducing conventional HIV testing uptake. We demonstrated that routinely collected programmatic data could be used to estimate the effects of HIVST kit distribution outside a trial environment. The methodology used in this paper could be replicated and implemented in different settings and enable more countries to routinely evaluate HIVST programming at the population level.

## Introduction

In 2019, up to 19% of people living with HIV (PLHIV) worldwide were not aware of their HIV status.^1^ In Western Africa, this proportion of undiagnosed PLHIV reached 33% in 2020.^2^ This is quite far from the *Joint United Nations Programme on HIV/AIDS* (UNAIDS) target to achieve less than 5% of PLHIV being undiagnosed by 2025. HIV testing is a crucial element of responses to HIV, as it is the first step to linkage to care and treatment. It is also key for prevention, as PLHIV who are on antiretroviral treatment (ART) and virally suppressed will not transmit HIV to their sexual partners.^3^

West African countries, including Côte d’Ivoire, are characterized by mixed epidemics: national HIV prevalence levels in the adult population are lower than those in East and Southern Africa (2·4% in Côte d’Ivoire in 2019 vs. 6·7% in East and Southern Africa), but HIV remains widespread, and higher prevalence levels are observed in key populations (KPs), such as female sex workers (FSW), men who have sex with men (MSM), and people who use drugs (PWUD) (between 3·4% and 12·3% in 2019).^4,5^ According to the UNAIDS, in West and Central Africa, 42% of new HIV infections (15-49 years old) in 2019 occurred among KPs and 27% among clients of sex workers and other sexual partners of this KP.^4^ A mathematical modeling study estimated that 44% of new infections in Côte d’Ivoire between 2005 and 2015 were due to partnerships between clients of FSW and their non-FSW female partners.^6^

The World Health Organization (WHO) recommends HIV self-testing (HIVST), which allows individuals to test and learn their results themselves when and how they want.^7^ It is an innovative tool that has been demonstrated to be safe, accurate, empowering, and acceptable and to also consistently increase the uptake and frequency of HIV testing across settings and populations.^8–15^ It is recommended that a reactive HIVST must be followed by a conventional test to confirm or disprove the results.

In Southern and East Africa, HIVST has been scaled up quickly, catalyzed by the Unitaid-funded Self-test Africa (STAR) initiative, which was started in 2015.^16^ However, before 2019, HIVST was offered only in West Africa through small-case pilot projects.^17^ A medium-scale HIVST program was implemented in Côte d’Ivoire, Mali, and Senegal in 2018, with an effective distribution of kits through the ATLAS project funded by Unitaid and implemented by a consortium led by *Solidarité thérapeutique et initiatives pour la santé* (Solthis) and the French Research Institute for Sustainable Development (IRD) since 2019.^18^ From 2019-2022, together with national programs, ATLAS planned to deliver 400 000 HIVST kits (214 000 in Côte d’Ivoire). The ATLAS program had set a target for 90% of HIVST implementation to reach KPs and their sexual partners, peers and clients. The remaining target of the HIVST implementation was to reach people with sexually transmitted infections (STIs), their partners and partners of people living with HIV.

ATLAS activities rely on both primary and secondary distribution channels. With primary distribution, HIVST kits are distributed by peer educators and frontline health care workers to primary contacts—MSM, PLHIV, STI patients, FSW, and PWUD—for their personal use. For secondary distribution, primary contacts are invited to redistribute HIVST kits to their peers, partners, clients and relatives (see Figure 1a). These secondary contacts are often members of key and vulnerable populations who often do not have easy access to the health system, including sexual partners of PLHIV or individuals in KPs. This specificity of HIVST kit distribution implies that HIVST beneficiaries (end users) are not limited to primary contacts. First, ATLAS’s program results have shown that HIVST can reduce stigma; preserve anonymity and confidentiality; reach KPs that do not access testing via other testing approaches; save opportunity costs for users and providers; empower users with autonomy and responsibility; and is noninvasive and easy to use.^19^

**Figure 1:**
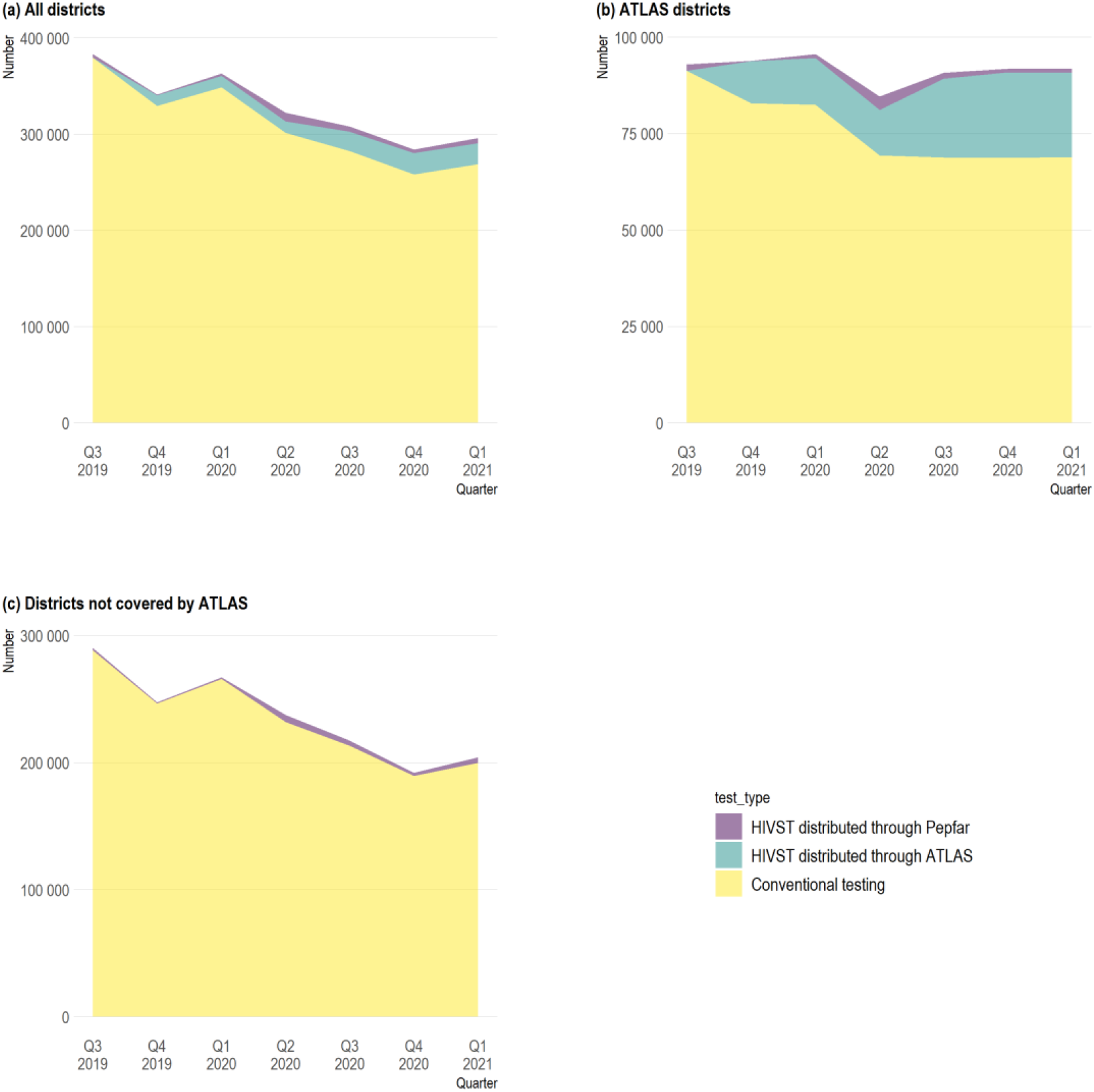
Number of conventional testing and HIVST kits distributed through PEPFAR and ATLAS from Q3 2019 to Q1 2021 in a) all 78 health districts monitored by PEPFAR in Côte d’Ivoire, b) the 9 ATLAS districts only, and c) the 69 districts not covered by ATLAS.

Several programs have developed methods to assess the use of HIVST and test results, such as supervision by health workers, the return of used kits, messages or phone call reminders to return used samples, the electronic transmission of photographs of test results, or the use of Bluetooth sensors.^20^ However, such tracking can be costly and counterproductive by limiting the use and distribution of HIVST and is not in line with the philosophy of HIVST, where users can anonymously decide when and where they are tested and if and to whom they want to report their results. The systematic tracking of HIVST though secondary distribution is logistically challenging and can also hinder the secondary distribution of HIVST, as primary contacts can be reluctant to redistribute an HIVST kit if they need to collect contact information. It could also be challenging for tracking HIVST at a large scale due to the logistics it might involve. To preserve the anonymity and confidentiality of those using HIVST and not impede the use of HIVST, ATLAS decided not to systematically track distributed HIVST kits. Nevertheless, HIVST users can still, if they wish, obtain additional support by calling a peer educator or a national HIV hotline.

Without systematic and direct feedback regarding the use and results of HIVST and linkage to confirmatory testing and ART, it is challenging to estimate the population-level impacts of HIVST distribution.^21^ In this paper, we aimed to circumvent this problem by using routinely collected programmatic data to estimate the effects of ATLAS’s HIVST distribution on access to HIV testing, conventional HIV testing (i.e., self-testing excluded), HIV diagnoses, and ART initiations in Côte d’Ivoire.

## Methods

### Data sources

In 2020, Côte d’Ivoire was divided into 33 health regions and 113 health districts.

ATLAS HIVST distribution in Côte d’Ivoire started during the third quarter of 2019 (Q3 2019) among individuals aged 16 years or older (minimum legal age for HIV testing without parental consent). All ATLAS implementing partners reported the number of HIVST kits distributed through ATLAS monthly by distribution site, delivery channel, age group and sex of primary contacts. Data were aggregated per health district and quarter of the year.

Routine programmatic data for adults over 15 years of age were obtained from the *President’s Emergency Plan for AIDS Relief (*PEPFAR) open-access public repository (https://data.pepfar.gov/). PEPFAR is the principal donor to the national AIDS program in Côte d’Ivoire. It collects programmatic data in the health districts where it intervenes, including (i) the number of HIVST kits distributed through PEPFAR-funded activities; (ii) conventional testing, i.e., the number of “individuals tested for HIV who received results”; (iii) HIV diagnoses, i.e., the number of “individuals who newly tested positive for HIV”; and (iv) ART initiations, i.e., the number of “people newly enrolled to receive ART”.

For this study, we used quarterly data aggregated at the health district level— harmonized according to the 2020 subdivision—from Q3 2019 to Q1 2021. Over this period, the PEPFAR data were only available for 78/113 (69%) Ivorian health districts. Only these districts were included in the analysis.

### Modeling strategy

The exact proportion of distributed HIVST kits that people actually use is unknown. We assumed, both for PEPFAR and ATLAS HIVST kits, a utilization rate (UR) of 80% based on the literature.^22^ We constructed a composite indicator reflecting access to HIV testing—all types—by summing the number of conventional tests and the number of HIVST kits that were actually used (UR 80%). We also considered a more conservative assumption with an HIVST UR of 60%.

Our analysis considered five outcomes: access to HIV testing (UR 80%), access to HIV testing (UR 60%), conventional testing, HIV diagnoses and ART initiations. The last three outcomes were obtained directly from the PEPFAR datasets.

We used ecological time series regression to model the linear effect of the number of HIVST kits distributed through ATLAS for each outcome.^23^ We first used linear mixed models with district-level random effects, as presented in Equation (1):

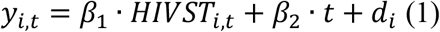

where *y*_*i,t*_ is the outcome of district *i* at time *t. HIVST* is the number of HIVST kits distributed through ATLAS. *β*_1_represents the effect of the latter variable on the outcomes. *t* is time, which captures conjectural effects. Conjectural effects (time) were modeled as a categorical variable of the quarter of the year to account for any nonlinear or nonpolynomial trend.^24^ *d*_*i*_ is the district-specific random effect. District-level random effects were used to account for autocorrelation due to multiple observations and to produce standard errors adjusted for clustering.

Then, contextual effects were also taken into account by introducing the categorical variable of health regions (*R*_*i*_) in the second model defined by Equation (2):

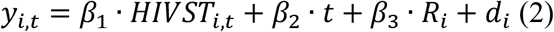

For each outcome, both Models (1) and (2) were run.

We performed the first sensitivity analysis by using cubic splines instead of a categorical variable for modeling time and compared the AIC (Akaike information criterion) of the models.

ATLAS activities were implemented in 9 of the 78 districts covered by the PEPFAR dataset. We also performed a second sensitivity analysis by restricting the sample to these 9 health districts.

All analyses were performed in R (version 4.0.3) using the lme4 package for statistical models.

## Results

### Descriptive statistics

In the 78 health districts monitored by PEPFAR, between Q3 2019 and Q1 2021, 30 781 HIVTS kits were distributed through PEPFAR, and 99 353 HIVST kits were distributed through ATLAS, compared with 2 167 828 conventional tests performed over the same period (Table 1). High disparities in terms of volume were observed between districts, with a minimum of 1 832 conventional tests and a maximum of 139 214 (median of 13 348). The 9 districts—out of 78—where ATLAS activities were implemented accounted for one quarter (532 287/2 167 828) of conventional tests. Important variations were also observed in terms of access to HIV testing, HIV diagnoses, and ART initiations across districts.

**Table 1:** District characteristics and activities between Q3 2019 and Q1 2021 in 78 health districts monitored by PEPFAR in Côte d’Ivoire

| Variable | All districts, N = 78 | ATLAS districts, N = 9 | Districts not covered by ATLAS, N = 69 |
| --- | --- | --- | --- |
| **HIV testing (HIVST UR: 80%)** |  |  |  |
| Total | 2 271 935 | 619 674 | 1 652 261 |
| Median | 19 417 | 71 044 | 18 351 |

| Variable | All districts, N =<br>78 | ATLAS districts, N<br>= 9 | Districts not covered by ATLAS,<br>N = 69 |
| --- | --- | --- | --- |
| Range | 1 834-149 762 | 15 087-149 762 | 1 834-80 050 |
| **HIV testing (HIVST UR:**<br>**60%)** |  |  |  |
| Total | 2 245 908 | 597 827 | 1 648 081 |
| Median | 19 399 | 68 315 | 18 304 |
| Range | 1 833-147 125 | 14 794-147 125 | 1 833-79 749 |
| **Conventional tests** |  |  |  |
| Total | 2 167 828 | 532 287 | 1 635 541 |
| Median | 19 348 | 57 037 | 18 162 |
| Range | 1 832-139 214 | 13 914-139 214 | 1 832-78 847 |
| **HIV diagnoses** |  |  |  |
| Total | 60 716 | 16 143 | 44 573 |
| Median | 484 | 1 465 | 467 |
| Range | 33-3 862 | 251-3 862 | 33-2 749 |
| **ART initiations** |  |  |  |
| Total | 54 354 | 13 846 | 40 508 |
| Median | 430 | 1 414 | 422 |
| Range | 33-3 068 | 216-3 068 | 33-2 274 |
| **HIVST kits distributed**<br>**through ATLAS** |  |  |  |
| Total | 99 353 | 99 353 | 0 |
| Median | 0 | 10 968 | 0 |
| Range | 0-23 472 | 1 364-23 472 | 0-0 |
| **HIVST kits distributed**<br>**through PEPFAR** |  |  |  |
| Total | 30 781 | 9 881 | 20 900 |
| Median | 168 | 735 | 100 |
| Range | 0-2 536 | 102-2 536 | 0-1 881 |

UR: utilization rate

In the 78 districts included in the analysis, conventional testing decreased between Q3 2019 and Q1 2021, from 379 554 individuals tested for HIV who received their results in Q3 2019 to 268 807 in Q1 2021 (Figure 1a). In the 69 districts that were not covered by ATLAS (Figure 1c), HIVST kits distributed through PEPFAR remained limited and largely insufficient to compensate for the reduction in conventional testing; only 13% of the tests in these districts were HIVST kits. In the 9 ATLAS districts, HIVST kit distribution—mainly through ATLAS, but also partially through PEPFAR—has increased continuously since Q3 2019 (Figure 1b), with a slow-down in Q2 2020, when governmental COVID-19 measures were introduced. Overall, HIVST in these districts mitigated the decreased access to HIV testing in these districts (Figure 2a & 2b).

**Figure 2:**
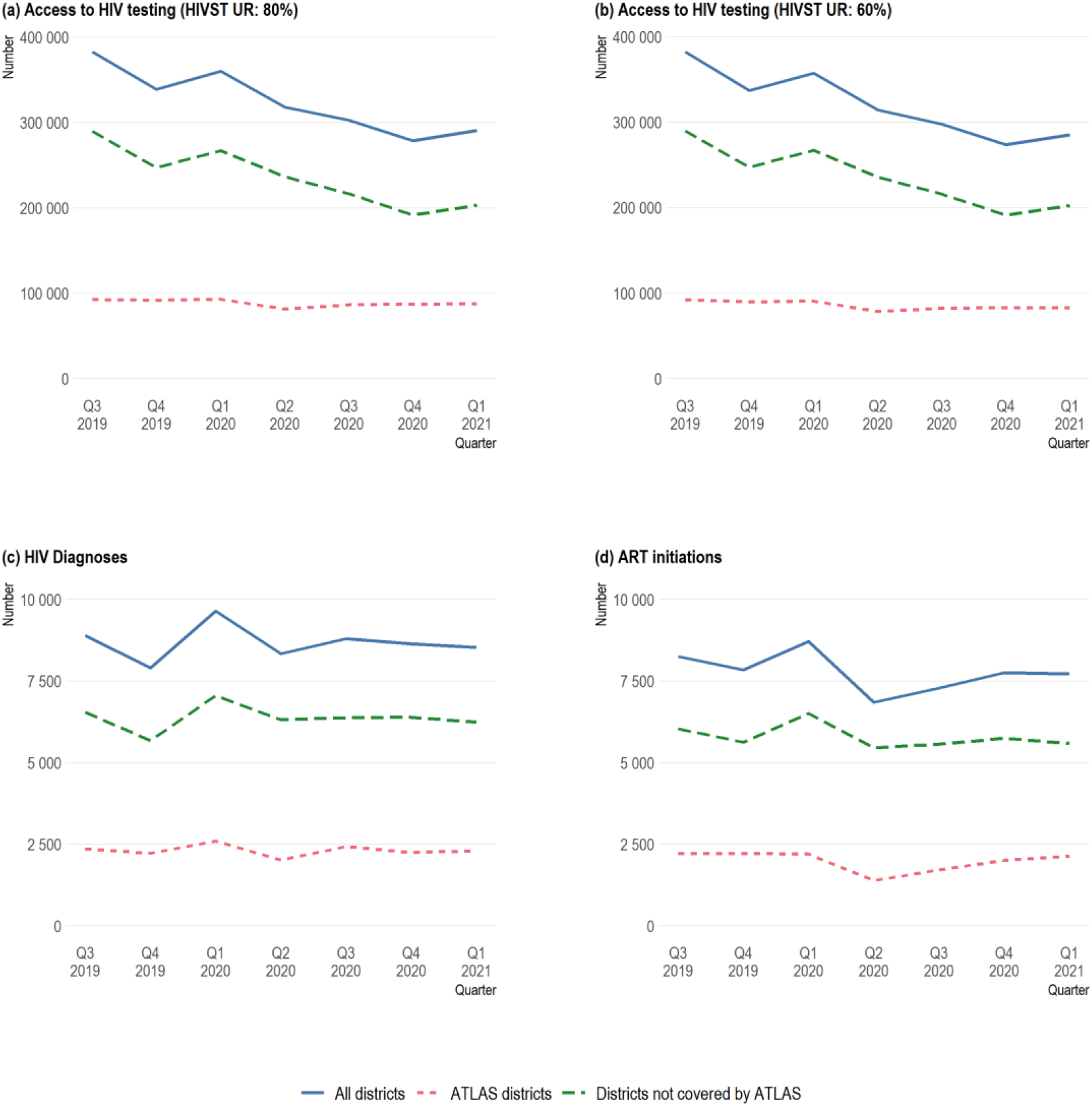
Number of testing, HIV diagnoses and ART initiations in the 78 health districts monitored by PEPFAR in Côte d’Ivoire (Q3 2019 to Q1 2021)

HIV diagnoses and ART initiations remained relatively stable over time (Figures 2c & 2d), with a catch-up effect observed in Q1 2020 after a slowdown in Q4 2019. Trends were fairly similar in the ATLAS districts and the districts not covered by ATLAS.

### Regression results

When adjusting for time and region, the model estimated a significant positive statistical relationship between ATLAS’s HIVST and access to HIV testing: for every 1000 additional HIVST kits distributed through ATLAS during a quarter in a district, ∼590 [95% confidence interval (CI): 357 to 821, p<0·001] additional individuals accessed HIV testing under the 80% UR hypothesis, and ∼390 [95% CI: 161 to 625, p<0·001] accessed testing under the 60% UR hypothesis. ATLAS HIVST kit distribution had a negative but non-statistically significant effect on conventional testing, with ∼-190 [95% CI: −427 to 37, p=0·10] conventional tests for every 1000 additional HIVST kits distributed by ATLAS. The association between HIVST kit distribution and HIV diagnoses was statistically significant and positive: +8 diagnoses [95% CI: 0 to 15, p=0·044]. No statistically significant association between HSVST kit distribution and ART initiations was observed: −2 [95% CI: −8 to 5, p=0·66]. Similar results were observed when adjusting only for time regarding the linear effect of the number of HIVST kits distributed through ATLAS on the different outcomes (first columns of Table 2).

**Table 2:** Linear effect of the number of HIVST kits distributed through ATLAS on access to HIV testing, conventional tests, diagnoses and ART initiations in the 78 health districts monitored by PEPFAR in Côte d’Ivoire (Q3 2019 to Q1 2021)

| Outcome | Adjusted for time |  |  | Adjusted for time and region |  |  |
| --- | --- | --- | --- | --- | --- | --- |
|  | Coef. | 95% CI<sup>1</sup> | P value | Coef. | 95% CI<sup>1</sup> | P value |
| HIV testing (HIVST UR: 80%) | 0.649 | 0.417 to 0.881 | <0.001 | 0.589 | 0.356 to 0.821 | <0.001 |
| HIV testing (HIVST UR: 60%) | 0.453 | 0.221 to 0.686 | <0.001 | 0.393 | 0.160 to 0.625 | <0.001 |
| Conventional tests | -0.133 | -0.365 to 0.099 | 0.26 | -0.195 | -0.427 to 0.038 | 0.10 |
| HIV diagnoses | 0.009 | 0.001 to 0.016 | 0.019 | 0.008 | 0.000 to 0.015 | 0.044 |
| ART initiations | 0.000 | -0.007 to 0.006 | 0.90 | -0.002 | -0.008 to 0.005 | 0.66 |

<sup>1</sup>CI = Confidence Interval. Coef.= coefficient. UR= utilization rate. For the five outcomes, only the regression coefficients of the number of HIVST kits distributed through ATLAS are presented. For the full regression table, see the Supplementary Material (Table A4 through A8).

The sensitivity analyses modeling time with cubic splines instead of categorical variables showed very similar results (Table A1) overall, except that the effect on HIV diagnoses was no longer significant at 5% (p=0·055). A comparison of the AIC values of the models indicated that the models with categorical variables fit the time series better (Table A2).

When restricting the analyses to the 9 ATLAS districts, the estimated magnitudes of association were larger. Only access to testing was statistically significantly associated with the number of HIVST kits distributed through ATLAS (Table A3).

## Discussion

Using routinely collected programmatic data aggregated quarterly at the health district level, our analysis showed a significant positive effect of HIVST kits distributed through ATLAS on access to HIV testing and HIV diagnoses, a negative but not significant effect on conventional testing and no observable effect on ART initiations.

HIVST could lead to some substitution effects if HIVST kits are used by individuals who would have undergone a conventional HIV test in its absence, as observed in other studies.^8,25^ Such effects may be concerning for policy-makers, as gains in HIV testing coverage due to HIVST distribution may result in a reduction in the number of conventional HIV tests. Our results did show a negative but nonsignificant effect of HIVST kit distribution on conventional testing (−195 conventional tests for every 1000 distributed HIVST), suggesting a possible but limited substitution effect. However, the overall net effect on HIV testing remained significantly positive, with +390 to +590 persons who would have not been tested otherwise, per 1000 distributed HIVST (depending on the assumption of the HIVST UR).

Moreover, the descriptive data showed that a decrease in conventional testing occurred in all districts, including those not covered by ATLAS activities, this is linked to the fact that PEPFAR’s testing strategies are revised annually and favoring more targeted approaches.^26^ Our results suggest that ATLAS HIVST distribution allowed to maintain access to HIV testing in its implementation districts despite the slowdown observed in Q2 2020 when governmental COVID-19 measures were introduced. In fact, the first results of the ATLAS project have shown that HIVST distribution activities among KPs could be easily adapted, including in the context of the COVID-19 pandemic.^27^

Due to its confidential nature, HIVST could overcome several structural barriers for HIV diagnoses—such as stigma and opportunity costs—and create new approaches to reach first-time testers and boost HIV retesting for KPs, therefore improving access to HIV testing overall. These results are in line with previous studies among KPs in East and South Africa.^8,25,28^

If HIVST is appropriately used as a triage test and individuals with reactive self-tests are linked for confirmatory testing, HIVST distribution activities should lead to a higher number of positive tests in conventional testing. Several actors have expressed concern that HIVST could have a negative impact on new diagnoses.^29^ Our results did not show any deleterious effect on HIV diagnoses but rather showed a significant positive effect. This is in line with some other studies, such as that by MacGowan *et al*., who found that the number of HIV infections detected in their HIVST arm was higher compared to the control arm in a randomized trial conducted among MSM.^30^

Our model did not observe any significant effect of ATLAS HIVST kit distribution on ART initiations. The estimated effect was negative when all 78 districts were included and positive when the analysis was limited to the 9 ATLAS districts. The analysis with 9 districts could suffer from a lack of power. A time effect could also be encountered if many newly diagnosed patients delayed ART initiation by several months; in such cases, the treatment initiation would not be registered and reported during the same quarter of the year.

The PEPFAR datasets are not exhaustive for Côte d’Ivoire and cover only 78 out of the 113 health districts at the national level and 9 of the 12 ATLAS districts.

Using aggregated data rather than individual data implies a lower number of observation points and therefore lower statistical power, although these data allow us to make population-level estimates. Aggregated data is subject to ecological bias and statistical relationships must be interpreted with caution. In addition, it is not possible to completely rule out any ‘contamination’ effect, as individuals living on district borders could perform conventional testing in the neighboring district. However, we could assume that population movement at boundaries could happen in both directions, thus compensating for each other, or expect the observed effect to have been even stronger without a ‘contamination’ effect. The collected data did not allow us to distinguish between confirmatory tests following HIVST and classic conventional tests, but as the HIV prevalence is relatively low in the country, the former number might not be important. Finally, HIVST kits distributed through UNICEF or the Global Fund to fight AIDS, tuberculosis and malaria were not taken into account when computing access to HIV testing, although the volumes of tests distributed by these programs were very low, 6 879 and 1 373 kits, respectively, by 2020, representing less than 7% of all HIVST kits distributed in the country.

A strength of this study is that it specifically used only indicators that had already been routinely collected by countries, which means that the method could be easily replicated in other contexts and used by other countries to monitor the impact of their HIVST activities without any additional cost. Our analysis did not rely on any systematic tracking system or data collection process, which can be expensive and complex and are counter to the rationale for HIVST.

The core component of the ATLAS HIVST strategy in West Africa is the secondary distribution of HIVST kits, primarily distributed through activities targeting individuals in KPs, in particular FSW and MSM. It is therefore expected that many HIVST users would not self-identify as being in a KP and that those with a reactive test would not link to partner community facilities serving KPs for confirmatory testing but rather to more general public or private facilities, making it difficult to link specific records with the distribution of HIVST kits. In addition, records of prior HIVST kit use at health facilities are expected to be underestimated, as recognizing such use would mean the individual was a member of and/or in a network of a KP. By using data aggregated at the district level and covering all testing facilities, confirmatory tests prompted by reactive HIVST results are considered, regardless of where they occurred. By allowing programs to shift from systematic tracking for evaluation, such indirect evaluation would help to focus on and increase access to HIV testing for hard-to-reach populations and first-time testers and allow large-scale secondary distribution implementation.

To the best of our knowledge, this represents the first study estimating the impact of HIVST kit distribution at the population level in West Africa. Such evaluation is pragmatic and could be performed with routinely collected programmatic indicators. The WHO recommends reporting on the number of HIVST kits distributed and estimating HIVST access and use through population-based surveys. Countries are burdened with multiple HIV reporting systems and numerous indicators. It could therefore be of considerable benefit to monitor the impact of self-testing through current data systems without introducing new indicators and further data collection. This method of triangulating available data provides further information on the population-level impacts of HIV self-testing to guide program use.

Our results highlight that a social network-based distribution strategy of HIVST, targeting key population members as primary contacts but aiming to reach their partners and social contacts, does have a positive impact on access to HIV testing and diagnoses that is observable at the population level.

## Data Availability

All data produced in the present study are available upon reasonable request to the authors

## Data sharing statements

Upon request to the first or last author

## Ethics statement

This study does not raise any ethical concern, as the data used are aggregated and thus completely anonymized.

A secondary analysis of the ATLAS programmatic data is included in the associated research protocol available at https://atlas.solthis.org/en/research/. This protocol (version 2.1, August 5 2019) was approved by the WHO Ethical Research Committee (August 7 2019, Reference: ERC 0003181), the National Ethics Committee for Life Sciences and Health of Côte d’Ivoire (May 28 2019, Reference: 049-19/MSHP/CNESVS-kp), the Ethics Committee of the Faculty of Medicine and Pharmacy of the University of Bamako, Mali (August 14 2019, Reference: 2019/88/CE/FMPOS), and the National Ethics Committee for Health Research of Senegal (July 26 2019, Protocol SEN19/32).

## Acknowledgments

The ATLAS Project was funded by Unitaid, Grant number 2018-23-ATLAS.

## Authors’ contribution

ASF and JL conceptualized the paper. ASF and PMD curated the data. ASF performed the formal analysis, wrote the initial computer programs and wrote the first draft of the paper, while JL supervised, validated and ensured the reproducibility of the results and was in charge of the project administration. ASF and JL defined the methodology and visualization adopted for the paper. ASF, PMD and AV provided the resources used for the research. JL, AV and CD secured funding for the project. ASF, KBK, EE, AV and PMD were involved in the investigation. All authors wrote, reviewed and edited the paper.

## Supplementary Material

### Supplementary figures

**Figure A1:**
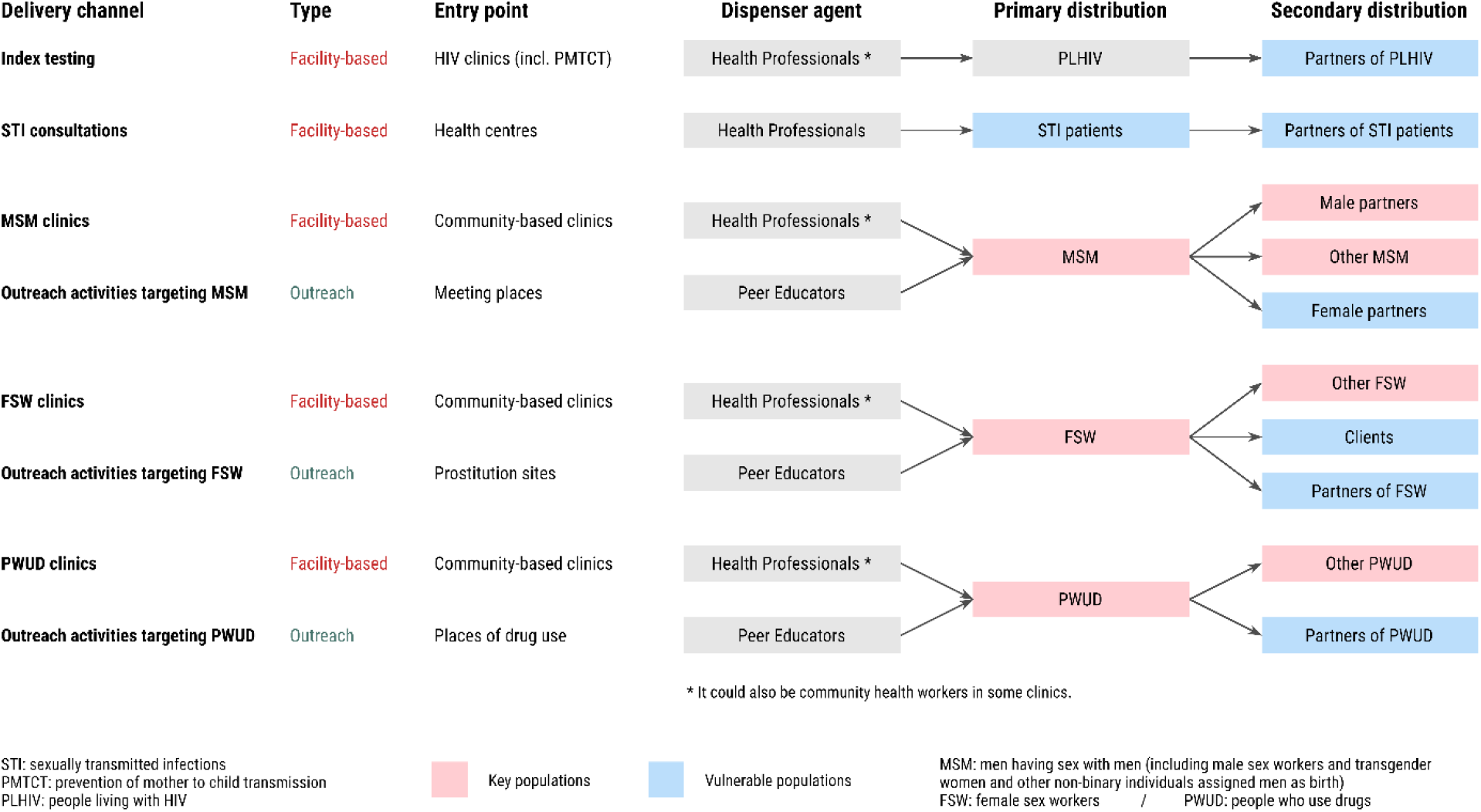
ATLAS HIVST kit delivery channels to reach key populations and other vulnerable populations. Note: This figure was first published in the ATLAS project protocol paper licensed under a Creative Commons Attribution.^1^

### Supplementary tables

**Table A1:**
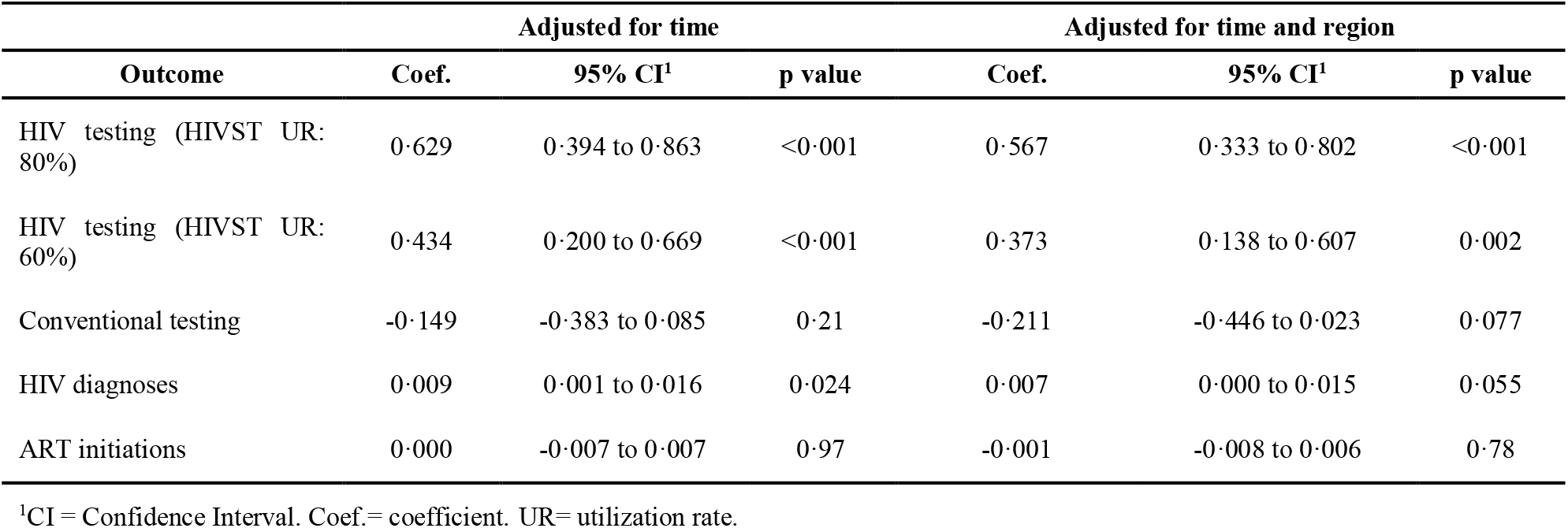
Effect of the number of HIVST kits distributed by ATLAS on HIV testing, conventional testing, diagnoses and ART initiations, time modeled using a cubic spline.

| Outcome | Adjusted for time |  |  | Adjusted for time and region |  |  |
| --- | --- | --- | --- | --- | --- | --- |
|  | Coef. | 95% CI<sup>1</sup> | p value | Coef. | 95% CI<sup>1</sup> | p value |
| HIV testing (HIVST UR: 80%) | 0.629 | 0.394 to 0.863 | <0.001 | 0.567 | 0.333 to 0.802 | <0.001 |
| HIV testing (HIVST UR: 60%) | 0.434 | 0.200 to 0.669 | <0.001 | 0.373 | 0.138 to 0.607 | 0.002 |
| Conventional testing | -0.149 | -0.383 to 0.085 | 0.21 | -0.211 | -0.446 to 0.023 | 0.077 |
| HIV diagnoses | 0.009 | 0.001 to 0.016 | 0.024 | 0.007 | 0.000 to 0.015 | 0.055 |
| ART initiations | 0.000 | -0.007 to 0.007 | 0.97 | -0.001 | -0.008 to 0.006 | 0.78 |

<sup>1</sup>CI = Confidence Interval. Coef.= coefficient. UR= utilization rate.

**Table A2:** Akaike information criterion comparison of the models according to trend modeling.

| Trend modeling | Outcome |  |  |  |  |
| --- | --- | --- | --- | --- | --- |
|  | HIV testing (HIVST UR: 80%) | HIV testing (HIVST UR: 60%) | Conventional testing | HIV diagnoses | ART initiations |
| Time indicator (categorical variable) | 8 908.4 | 8 907.5 | 8 906.9 | 5 365.0 | 5 264.5 |
| Cubic splines | 8 949.9 | 8 948.7 | 8 947.4 | 5 393.5 | 5 293.2 |

Note: AIC was compared for the models adjusted for time and region. UR= utilization rate.

**Table A3:** Effect of the number of HIVST kits distributed by ATLAS on HIV testing, conventional testing, diagnoses and ART initiations, data restricted to the ATLAS districts.

| Outcome | Adjusted for time |  |  | Adjusted for time and region |  |  |
| --- | --- | --- | --- | --- | --- | --- |
|  | Coef. | 95% CI<sup>1</sup> | p value | Coef. | 95% CI<sup>1</sup> | p value |
| HIV testing (HIVST UR: 80%) | 0.997 | 0.357 to 1.637 | 0.003 | 0.910 | 0.265 to 1.554 | 0.007 |
| HIV testing (HIVST UR: 60%) | 0.798 | 0.160 to 1.436 | 0.015 | 0.710 | 0.068 to 1.353 | 0.031 |
| Conventional testing | 0.199 | -0.435 to 0.833 | 0.53 | 0.112 | -0.527 to 0.750 | 0.73 |
| HIV diagnoses | 0.018 | -0.008 to 0.043 | 0.17 | 0.014 | -0.010 to 0.038 | 0.25 |
| ART initiations | 0.004 | -0.016 to 0.024 | 0.70 | 0.005 | -0.014 to 0.025 | 0.57 |

<sup>1</sup>CI = Confidence Interval. Coef.= coefficient. UR= utilization rate. Time is modeled as a categorical variable.

**Table A4.** Linear effect of the number of HIVST kits distributed through ATLAS on access to HIV testing (HIVST UR: 80%) in the 78 health districts monitored by PEPFAR in Côte d’Ivoire (Q3 2019 to Q1 2021)

| Outcome | Adjusted for time |  |  | Adjusted for time and region |  |  |
| --- | --- | --- | --- | --- | --- | --- |
|  | Coef. | 95% CI<sup>1</sup> | p value | Coef. | 95% CI<sup>1</sup> | p value |
| Number of HIVST kits distributed through ATLAS | 0.649 | 0.417 to 0.881 | <0.001 | 0.589 | 0.356 to 0.821 | <0.001 |
| Year, quarter |  |  |  |  |  |  |
| 2019 Q3 | — | — |  | — | — |  |
| 2019 Q4 | -648.483 | -949.095 to -347.872 | <0.001 | -640.099 | -940.512 to -339.686 | <0.001 |
| 2020 Q1 | -388.826 | -689.875 to -87.777 | 0.011 | -379.439 | -680.290 to -78.587 | 0.014 |
| 2020 Q2 | -923.909 | -1 224.827 to -622.991 | <0.001 | -914.811 | -1 215.531 to -614.091 | <0.001 |
| 2020 Q3 | -1 191.344 | -1 496.342 to -886.346 | <0.001 | -1 175.526 | -1 480.335 to -870.717 | <0.001 |
| 2020 Q4 | -1 514.979 | -1 821.074 to -1 208.883 | <0.001 | -1 497.785 | -1 803.694 to -1 191.876 | <0.001 |
| 2021 Q1 | -1 360.697 | -1 666.589 to -1 054.804 | <0.001 | -1 343.749 | -1 649.455 to -1 038.044 | <0.001 |
| Health region |  |  |  |  |  |  |
| Abidjan 1-Grands Ponts | — | — |  | — | — |  |
| Abidjan 2 |  |  |  | 6 277.155 | 2 830.228 to 9 724.082 | <0.001 |
| Agneby-Tiassa |  |  |  | -2 671.299 | -6 479.104 to 1 136.505 | 0.17 |
| Bagoue |  |  |  | -4 839.861 | -9 286.027 to -393.695 | 0.033 |
| Belier |  |  |  | -2 804.795 | -6 612.600 to 1 003.010 | 0.15 |
| Bere |  |  |  | -4 984.623 | -8 792.428 to -1 176.818 | 0.011 |
| Cavally |  |  |  | -4 840.133 | -9 286.299 to -393.967 | 0.033 |
| Gbeke |  |  |  | -2 734.833 | -5 772.984 to 303.319 | 0.077 |
| Gbokle-Nawa-San Pedro |  |  |  | 1 239.698 | -2 210.768 to 4 690.163 | 0.47 |
| Goh |  |  |  | -2 497.642 | -6 305.447 to 1 310.163 | 0.19 |
| Gontougo |  |  |  | -3 941.187 | -7 148.081 to -734.293 | 0.017 |
| Guemon |  |  |  | -3 694.680 | -7 502.485 to 113.124 | 0.057 |
| Hambol |  |  |  | -3 723.804 | -9 688.319 to 2 240.711 | 0.22 |
| Haut-Sassandra |  |  |  | -1 280.947 | -4 725.489 to 2 163.595 | 0.46 |
| Iffou |  |  |  | -3 694.718 | -9 659.233 to 2 269.796 | 0.22 |
| Indenie-Djuablin |  |  |  | -2 107.190 | -6 553.356 to 2 338.976 | 0.35 |
| Loh-Djiboua |  |  |  | 317.224 | -5 647.290 to 6 281.739 | 0.92 |
| Marahoue |  |  |  | -1 896.518 | -5 704.323 to 1 911.286 | 0.32 |
| Me |  |  |  | -3 430.036 | -6 873.947 to 13.875 | 0.051 |
|  | Coef. | 95% CI<sup>1</sup> | p value | Coef. | 95% CI<sup>1</sup> | p value |
| Moronou |  |  |  | -4 413.957 | -8 221.761 to -606.152 | 0.024 |
| N'zi |  |  |  | -5 155.861 | -8 963.666 to -1 348.056 | 0.009 |
| Poro |  |  |  | -3 401.490 | -7 209.295 to 406.315 | 0.079 |
| Tchologo |  |  |  | -5 147.618 | -9 593.785 to -701.452 | 0.024 |
| Tonkpi |  |  |  | -3 196.842 | -7 004.647 to 610.963 | 0.10 |
| Worodougou |  |  |  | -3 398.676 | -7 844.842 to 1 047.490 | 0.13 |

<sup>1</sup>CI = Confidence Interval. Coef.= coefficient.

**Table A5.** Linear effect of the number of HIVST kits distributed through ATLAS on access to HIV testing (HIVST UR: 60%) in the 78 health districts monitored by PEPFAR in Côte d’Ivoire (Q3 2019 to Q1 2021)

| Outcome | Adjusted for time |  |  | Adjusted for time and region |  |  |
| --- | --- | --- | --- | --- | --- | --- |
|  | Coef. | 95% CI<sup>1</sup> | p value | Coef. | 95% CI<sup>1</sup> | p value |
| Number of HIVST kits distributed through ATLAS | 0.453 | 0.221 to 0.686 | <0.001 | 0.393 | 0.160 to 0.625 | <0.001 |
| Year, quarter |  |  |  |  |  |  |
| 2019 Q3 | — | — |  | — | — |  |
| 2019 Q4 | -642.300 | -942.751 to -341.850 | <0.001 | -633.892 | -934.143 to -333.641 | <0.001 |
| 2020 Q1 | -385.996 | -686.884 to -85.109 | 0.012 | -376.582 | -677.271 to -75.893 | 0.014 |
| 2020 Q2 | -938.792 | -1 239.549 to -638.036 | <0.001 | -929.668 | -1 230.226 to -629.110 | <0.001 |
| 2020 Q3 | -1 196.979 | -1 501.813 to -892.146 | <0.001 | -1 181.116 | -1 485.760 to -876.472 | <0.001 |
| 2020 Q4 | -1 515.986 | -1 821.916 to -1 210.057 | <0.001 | -1 498.744 | -1 804.486 to -1 193.001 | <0.001 |
| 2021 Q1 | -1 366.110 | -1 671.837 to -1 060.383 | <0.001 | -1 349.114 | -1 654.654 to -1 043.575 | <0.001 |
| Health region |  |  |  |  |  |  |
| Abidjan 1-Grands Ponts | — | — |  | — | — |  |
| Abidjan 2 |  |  |  | 6 251.295 | 2 818.041 to 9 684.549 | <0.001 |
| Agneby-Tiassa |  |  |  | -2 655.936 | -6 448.617 to 1 136.745 | 0.17 |
| Bagoue |  |  |  | -4 818.322 | -9 246.826 to -389.817 | 0.034 |
| Belier |  |  |  | -2 790.117 | -6 582.798 to 1 002.564 | 0.15 |
| Bere |  |  |  | -4 963.298 | -8 755.979 to -1 170.617 | 0.011 |
| Cavally |  |  |  | -4 823.365 | -9 251.869 to -394.860 | 0.033 |
| Gbeke |  |  |  | -2 722.646 | -5 748.734 to 303.443 | 0.077 |
| Gbokle-Nawa-San Pedro |  |  |  | 1 222.206 | -2 214.597 to 4 659.008 | 0.48 |
| Goh |  |  |  | -2 486.384 | -6 279.065 to 1 306.297 | 0.19 |
|  | Coef. | 95% CI<sup>1</sup> | p value | Coef. | 95% CI<sup>1</sup> | p value |
| Gontougo |  |  |  | -3 921·359 | -7 115·518 to -727·199 | 0·017 |
| Guemon |  |  |  | -3 682·555 | -7 475·236 to 110·126 | 0·057 |
| Hambol |  |  |  | -3 704·422 | -9 645·239 to 2 236·396 | 0·22 |
| Haut-Sassandra |  |  |  | -1 270·208 | -4 701·070 to 2 160·655 | 0·46 |
| Iffou |  |  |  | -3 673·822 | -9 614·639 to 2 266·996 | 0·22 |
| Indenie-Djuablin |  |  |  | -2 092·336 | -6 520·840 to 2 336·168 | 0·35 |
| Loh-Djiboua |  |  |  | 327·207 | -5 613·611 to 6 268·024 | 0·91 |
| Marahoue |  |  |  | -1 892·708 | -5 685·389 to 1 899·974 | 0·32 |
| Me |  |  |  | -3 419·005 | -6 849·235 to 11·226 | 0·051 |
| Moronou |  |  |  | -4 394·060 | -8 186·741 to -601·379 | 0·024 |
| N'zi |  |  |  | -5 134·441 | -8 927·122 to -1 341·760 | 0·009 |
| Poro |  |  |  | -3 382·650 | -7 175·332 to 410·031 | 0·079 |
| Tchologo |  |  |  | -5 126·408 | -9 554·912 to -697·903 | 0·024 |
| Tonkpi |  |  |  | -3 186·260 | -6 978·941 to 606·421 | 0·10 |
| Worodougou |  |  |  | -3 376·808 | -7 805·312 to 1 051·697 | 0·13 |

<sup>1</sup>CI = Confidence Interval. Coef.= coefficient.

**Table A6.**
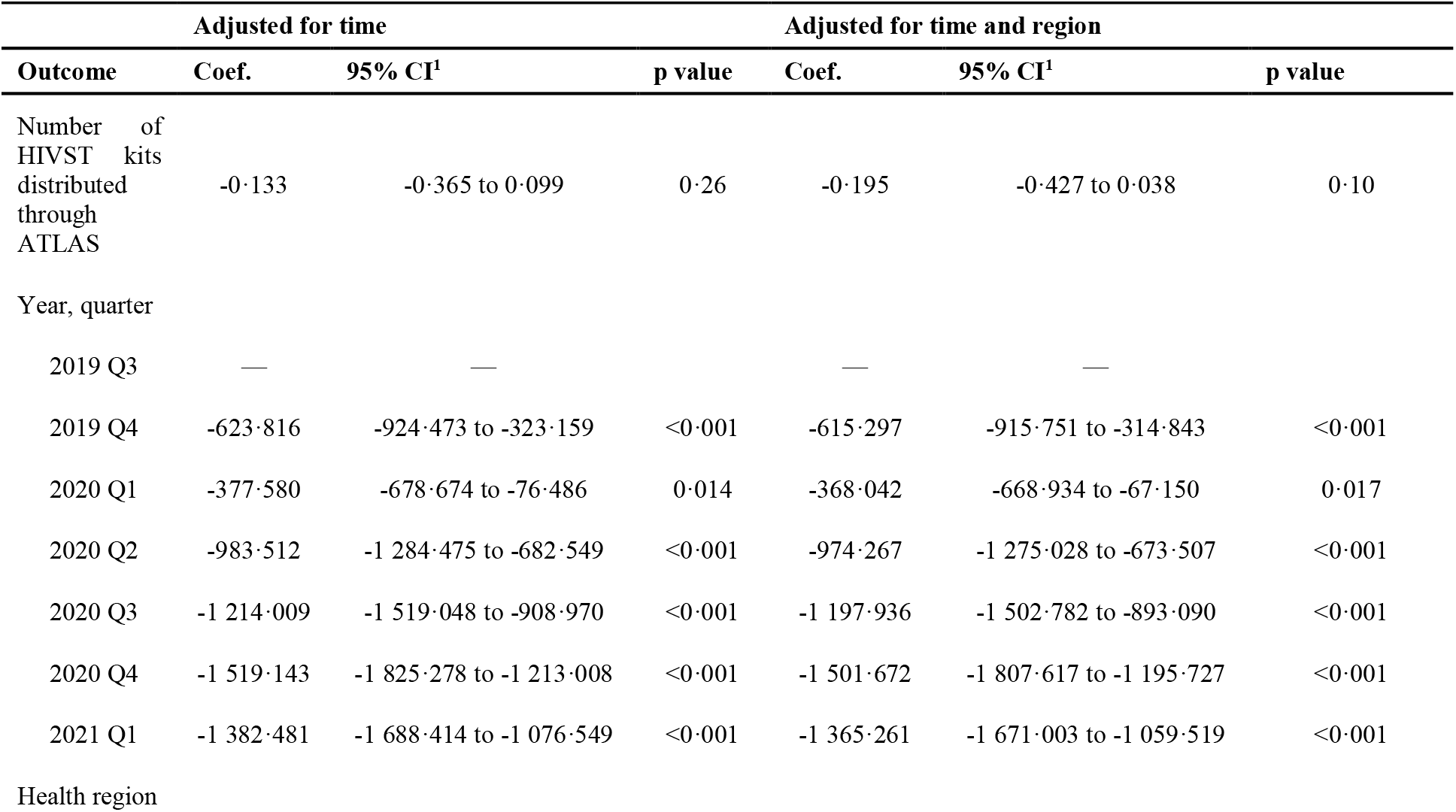
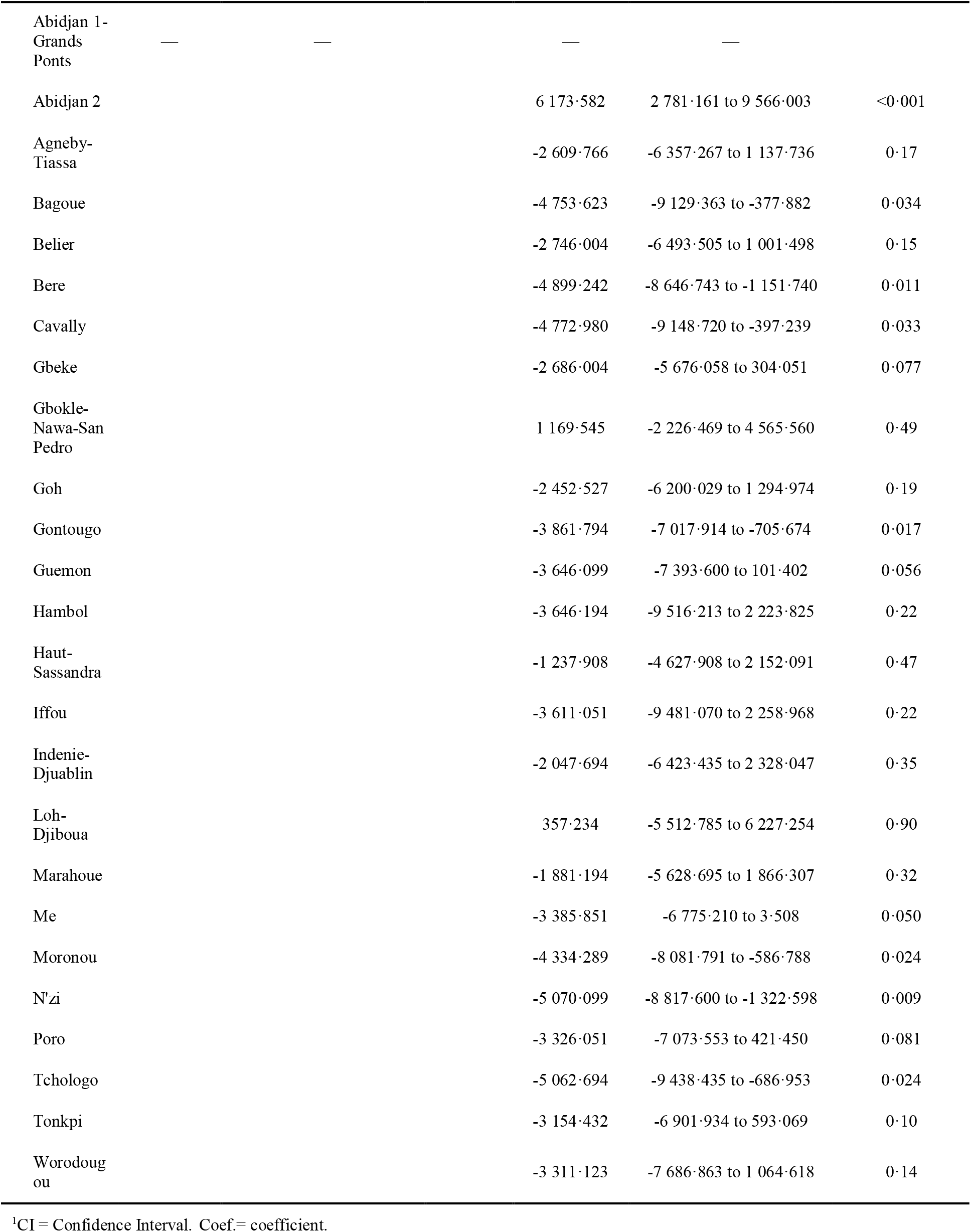
Linear effect of the number of HIVST kits distributed through ATLAS on HIV conventional testing in the 78 health districts monitored by PEPFAR in Côte d’Ivoire (Q3 2019 to Q1 2021)

**Table A7.** Linear effect of the number of HIVST kits distributed through ATLAS on HIV diagnoses in the 78 health districts monitored by PEPFAR in Côte d’Ivoire (Q3 2019 to Q1 2021)

|  |  | Adjusted for time |  |  | Adjusted for time and region |  |  |
| --- | --- | --- | --- | --- | --- | --- | --- |
| Outcome |  | Coef. | 95% CI<sup>1</sup> | p value | Coef. | 95% CI<sup>1</sup> | p value |
| Number of HIVST kits distributed through ATLAS |  | 0.009 | 0.001 to 0.016 | 0.019 | 0.008 | 0.000 to 0.015 | 0.044 |
| Year, quarter |  |  |  |  |  |  |  |
| 2019 Q3 |  | — | — |  | — | — |  |
| 2019 Q4 |  | -14.016 | -23.595 to -4.437 | 0.004 | -13.838 | -23.413 to -4.263 | 0.005 |
| 2020 Q1 |  | 8.208 | -1.385 to 17.801 | 0.093 | 8.407 | -1.182 to 17.996 | 0.086 |
| 2020 Q2 |  | -8.557 | -18.146 to 1.032 | 0.080 | -8.364 | -17.949 to 1.220 | 0.087 |
| 2020 Q3 |  | -3.623 | -13.342 to 6.096 | 0.46 | -3.288 | -13.003 to 6.427 | 0.51 |
| 2020 Q4 |  | -5.851 | -15.605 to 3.903 | 0.24 | -5.487 | -15.237 to 4.263 | 0.27 |
| 2021 Q1 |  | -7.238 | -16.986 to 2.510 | 0.15 | -6.879 | -16.623 to 2.865 | 0.17 |
| Health region |  |  |  |  |  |  |  |
| Abidjan 1-Grands Ponts |  | — | — |  | — | — |  |
| Abidjan 2 |  |  |  |  | 210.178 | 101.255 to 319.100 | <0.001 |
| Agneby-Tiassa |  |  |  |  | -128.466 | -248.790 to -8.142 | 0.037 |
| Bagoue |  |  |  |  | -160.990 | -301.486 to -20.494 | 0.026 |
| Belier |  |  |  |  | -115.561 | -235.886 to 4.763 | 0.059 |
| Bere |  |  |  |  | -167.371 | -287.695 to -47.047 | 0.007 |
| Cavally |  |  |  |  | -142.418 | -282.914 to -1.923 | 0.047 |
| Gbeke |  |  |  |  | -109.061 | -205.065 to -13.057 | 0.027 |
| Gbokle-Nawa-San Pedro |  |  |  |  | -78.665 | -187.701 to 30.371 | 0.15 |
| Goh |  |  |  |  | -85.466 | -205.790 to 34.858 | 0.16 |
| Gontougo |  |  |  |  | -122.990 | -224.326 to -21.654 | 0.018 |
| Guemon |  |  |  |  | -126.228 | -246.552 to -5.904 | 0.040 |
| Hambol |  |  |  |  | -132.133 | -320.607 to 56.342 | 0.17 |
| Haut-Sassandra |  |  |  |  | -79.490 | -188.335 to 29.356 | 0.15 |
| Iffou |  |  |  |  | -144.847 | -333.321 to 43.627 | 0.13 |
| Indenie-Djuablin |  |  |  |  | -55.776 | -196.271 to 84.720 | 0.43 |
| Loh-Djiboua |  |  |  |  | -39.561 | -228.036 to 148.913 | 0.68 |
| Marahoue |  |  |  |  | -97.704 | -218.028 to 22.620 | 0.11 |
| Me |  |  |  |  | -156.063 | -264.888 to -47.238 | 0.006 |

| Outcome | Adjusted for time |  |  | Adjusted for time and region |  |  |
| --- | --- | --- | --- | --- | --- | --- |
|  | Coef. | 95% CI<sup>1</sup> | p value | Coef. | 95% CI<sup>1</sup> | p value |
| Moronou |  |  |  | -162.228 | -282.552 to -41.904 | 0.009 |
| N'zi |  |  |  | -180.895 | -301.219 to -60.570 | 0.004 |
| Poro |  |  |  | -116.799 | -237.124 to 3.525 | 0.057 |
| Tchologo |  |  |  | -169.276 | -309.771 to -28.780 | 0.019 |
| Tonkpi |  |  |  | -81.466 | -201.790 to 38.858 | 0.18 |
| Worodougou |  |  |  | -149.776 | -290.271 to -9.280 | 0.037 |

<sup>1</sup>CI = Confidence Interval. Coef.= coefficient.

**Table A8.** Linear effect of the number of HIVST kits distributed through ATLAS on ART initiations in the 78 health districts monitored by PEPFAR in Côte d’Ivoire (Q3 2019 to Q1 2021)

| Outcome | Adjusted for time |  |  | Adjusted for time and region |  |  |
| --- | --- | --- | --- | --- | --- | --- |
|  | Coef. | 95% CI<sup>1</sup> | p value | Coef. | 95% CI<sup>1</sup> | p value |
| Number of HIVST kits distributed through ATLAS | 0.000 | -0.007 to 0.006 | 0.90 | -0.002 | -0.008 to 0.005 | 0.66 |
| Year, quarter |  |  |  |  |  |  |
| 2019 Q3 | — | — |  | — | — |  |
| 2019 Q4 | -5.220 | -13.918 to 3.477 | 0.24 | -5.073 | -13.766 to 3.620 | 0.25 |
| 2020 Q1 | 6.031 | -2.679 to 14.741 | 0.17 | 6.196 | -2.510 to 14.902 | 0.16 |
| 2020 Q2 | -17.946 | -26.652 to -9.240 | <0.001 | -17.786 | -26.488 to -9.084 | <0.001 |
| 2020 Q3 | -12.307 | -21.131 to -3.482 | 0.006 | -12.028 | -20.849 to -3.208 | 0.008 |
| 2020 Q4 | -6.181 | -15.037 to 2.675 | 0.17 | -5.879 | -14.731 to 2.973 | 0.19 |
| 2021 Q1 | -6.632 | -15.482 to 2.219 | 0.14 | -6.334 | -15.180 to 2.513 | 0.16 |
| Health region |  |  |  |  |  |  |
| Abidjan 1-Grands Ponts | — | — |  | — | — |  |
| Abidjan 2 |  |  |  | 179.593 | 81.829 to 277.356 | <0.001 |
| Agneby-Tiassa |  |  |  | -111.742 | -219.738 to -3.746 | 0.043 |
| Bagoue |  |  |  | -143.313 | -269.414 to -17.213 | 0.027 |
| Belier |  |  |  | -105.599 | -213.595 to 2.397 | 0.055 |
| Bere |  |  |  | -151.789 | -259.785 to -43.794 | 0.007 |
| Cavally |  |  |  | -147.956 | -274.056 to -21.856 | 0.022 |
| Gbeke |  |  |  | -99.242 | -185.410 to -13.074 | 0.025 |
| Gbokle-Nawa-San Pedro |  |  |  | -71.021 | -168.889 to 26.846 | 0.15 |
|  | Coef. | 95% CI<sup>1</sup> | p value | Coef. | 95% CI<sup>1</sup> | p value |
| Goh |  |  |  | -83.170 | -191.166 to 24.825 | 0.13 |
| Gontougo |  |  |  | -109.970 | -200.924 to -19.017 | 0.019 |
| Guemon |  |  |  | -131.361 | -239.357 to -23.365 | 0.018 |
| Hambol |  |  |  | -120.885 | -290.047 to 48.278 | 0.16 |
| Haut-Sassandra |  |  |  | -64.028 | -161.721 to 33.666 | 0.19 |
| Iffou |  |  |  | -129.885 | -299.047 to 39.278 | 0.13 |
| Indenie-Djuablin |  |  |  | -40.028 | -166.128 to 86.073 | 0.53 |
| Loh-Djiboua |  |  |  | -38.170 | -207.333 to 130.992 | 0.65 |
| Marahoue |  |  |  | -86.647 | -194.642 to 21.349 | 0.11 |
| Me |  |  |  | -142.715 | -240.389 to -45.040 | 0.005 |
| Moronou |  |  |  | -144.789 | -252.785 to -36.794 | 0.010 |
| N'zi |  |  |  | -163.361 | -271.357 to -55.365 | 0.004 |
| Poro |  |  |  | -105.789 | -213.785 to 2.206 | 0.055 |
| Tchologo |  |  |  | -149.742 | -275.842 to -23.642 | 0.021 |
| Tonkpi |  |  |  | -83.123 | -191.118 to 24.873 | 0.13 |
| Worodougou |  |  |  | -127.242 | -253.342 to -1.142 | 0.048 |

<sup>1</sup>CI = Confidence Interval. Coef.= coefficient.

### Composition of the ATLAS Team

| **ATLAS Research Team** |  |
| --- | --- |
| Amani Elvis Georges | Programme PACCI, ANRS Research Site, Treichville University Hospital, Abidjan, Côte d'Ivoire. |
| Badiane Kéba | Solthis, Sénégal |
| Bayac Céline | Solthis, France |
| Bekelynck Anne | Programme PACCI, ANRS Research Site, Treichville University Hospital, Abidjan, Côte d'Ivoire |
| Boily Marie-Claude | Medical Research Council Centre for Global Infectious Disease Analysis, Department of Infectious Disease Epidemiology, Imperial College London, London, United Kingdom |
| Boye Sokhna | Centre Population et Développement, Institut de Recherche pour le Développement, Université de Paris, Inserm, Paris, France |
| Breton Guillaume | Solthis, Paris, France |
| d'Elbée Marc | Department of Global Health and Development, Faculty of Public Health and Policy, London School of Hygiene and Tropical Medicine, London, UK |
| Desclaux Alice | Institut de Recherche pour le Développement, Transvihmi (UMI 233 IRD, 1175 INSERM, Montpellier University), Montpellier, France/CRCF, Dakar, Sénégal |
| Desgrées du Loû Annabel | Centre Population et Développement, Institut de Recherche pour le Développement, Université de Paris, Inserm, Paris, France |
| Diop Papa Moussa | Solthis, Sénégal |
| Doumenc-Aïdara Clémence | Solthis, Dakar, Sénégal |
| Ehui Eboi | Directeur Coordonnateur, PNLS |
| Graham Medley | Department of Global Health and Development, Faculty of Public Health and Policy, London School of Hygiene and Tropical Medicine, London, UK |
| Jean Kévin | Laboratoire MESuRS, Conservatoire National des Arts et Métiers, Paris, France |
| Keita Abdelaye | Institut National de Recherche en Santé Publique, Bamako, Mali |
| Kouadio Brou Alexis | Assistant de recherche, Côte d'Ivoire |
| Kouassi Kra Arsène | Centre Population et Développement, Institut de Recherche pour le Développement, Université de Paris, Inserm, Paris, France |
| Ky-Zerbo Odette | TransVIHMI, IRD, Université de Montpellier, INSERM |
| Larmarange Joseph | Centre Population et Développement, Institut de Recherche pour le Développement, Université de Paris, Inserm, Paris, France |
| Maheu-Giroux Mathieu | Department of Epidemiology, Biostatistics, and Occupational Health, School of Population and Global Health, McGill University, Montréal, QC, H3A 1A2, Canada |
| Medley Graham | Department of Global Health and Development, Faculty of Public Health and Policy, London School of Hygiene and Tropical Medicine, London, UK |
| Moh Raoul | Programme PACCI, ANRS Research Site, Treichville University Hospital, Abidjan, Côte d'Ivoire.<br>Department of Infectious and Tropical Diseases, Treichville University Teaching Hospital, Abidjan, Côte d'Ivoire.<br>Medical School, University Felix Houphouet Boigny, Abidjan, Côte d'Ivoire |
| Ndour Cheikh Tidiane | Division de Lutte contre le Sida et les IST, Ministère de la Santé et de l'Action Sociale Institut d'Hygiène Sociale, Dakar, Sénégal |
| Pourette Dolorès | Centre Population et Développement, Institut de Recherche pour le Développement, Université de Paris, Inserm, Paris, France |
| Rouveau Nicolas | Centre Population et Développement, Institut de Recherche pour le Développement, Université de Paris, Inserm, Paris, France |
| Silhol Romain | Medical Research Council Centre for Global Infectious Disease Analysis, Department of Infectious Disease Epidemiology, Imperial College London, London, United Kingdom |
| Simo Fotso Arlette | Centre Population et Développement, Institut de Recherche pour le Développement, Université de Paris, Inserm, Paris, France |
| Terris-Prestholt Fern | Department of Global Health and Development, Faculty of Public Health and Policy, London School of Hygiene and Tropical Medicine, London, UK |
| Traore Métogara Mohamed | Solthis, Côte d'Ivoire |
| Vautier Anthony | Solthis, Dakar, Sénégal |
| Solthis coordination team |  |
| Diallo Sanata | Solthis, Dakar, Sénégal |
| Gueye Papa Alioune | Solthis, Dakar, Sénégal |
| Geoffroy Olivier | Solthis, Abidjan, Côte d'Ivoire |
| Kabemba Odé Kanku | Solthis, Bamako, Mali |
| Implementation in Côte d'Ivoire |  |
| Abokon Armand | Fondation Ariel Glaser, Côte d'Ivoire |
| Anoma Camille | Espace Confiance, Côte d'Ivoire |
| Diokouri Annie | Fondation Ariel Glaser, Côte d'Ivoire |
| Kouame Blaise | Service Dépistage, PNLS |
| Kouakou Venance | Heartland Alliance, Côte d'Ivoire |
| Koffi Odette | Aprosam, Côte d'Ivoire |
| Kpolo Alain-Michel | Ruban Rouge, Côte d'Ivoire |
| Tety Josiane | Blety, Côte d'Ivoire |
| Traore Yacouba | ORASUR, Côte d'Ivoire |
| Implementation in Mali |  |
| Bagendabanga Jules | FHI 360, Mali |
| Berthé Djelika | PSI, Mali |
| Diakite Daouda | Secrétariat Exécutif du Haut Conseil National de Lutte contre le Sida, Mali |
| Diakité Mahamadou | Danayaso, Mali |
| Diallo Youssouf | CSLS/MSHP |
| Daouda Minta | Comité scientifique VIH |
| Hessou Septime | Plan Mali |
| Kanambaye Saidou | PSI, Mali |
| Kanoute Abdul Karim | Plan Mali |
| Keita Dembele Bintou | Arcad-Sida, Mali |
| Koné Dramane | Secrétariat Exécutif du Haut Conseil National de Lutte contre le Sida, Mali |
| Koné Mariam | AKS, Mali |
| Maiga Almoustapha | Comité scientifique VIH |
| Nouhoum Telly | CSLS/MSHP |
| Sanogo Abdoulaye | Amprode Sahel, Mali |
| Saran Keita Aminata | Soutoura, Mali |
| Sidibé Fadiala | Soutoura, Mali |
| Tall Madani | FHI 360, Mali |
| Yattassaye Camara Adam | Arcad-Sida, Mali |
| Implementation in Senegal |  |
| Bâ Idrissa | CEPIAD, Sénégal |
| Diallo Papa Amadou Niang | CNLS, Sénégal |
| Fall Fatou | DLSI, Ministère de la Santé et de l'action sociale, Sénégal |
| Guèye NDèye Fatou NGom | CTA, Sénégal |
| Ndiaye Sidy Mokhtar | Enda Santé, Sénégal |
| Niang Alassane Moussa | DLSI, Ministère de la Santé et de l'action sociale, Sénégal |
| Samba Oumar | CEPIAD, Sénégal |
| Thiam Safiatou | CNLS, Sénégal |
| Turpin Nguissali M.E. | Enda Santé, Sénégal |
| Partners |  |
| Bouaré Seydou | Assistant de recherche, Mali |
| Camara Cheick Sidi | Assistant de recherche, Mali |
| Eponon Ehua Agnes | Assistante de recherche, Sénégal |
| Kouvahe Amélé | Stagiaire IRD |
| Montaufray Marie-Anne | Stagiaire IRD |
| Mosso Rosine | ENSEA Ecole Nationale de Statistiques et d'Economie Appliquée, Abidjan, Côte d'Ivoire |
| Ndeye Pauline Dama | Chargée de l'enquête coupons, Sénégal |
| Sarrassat Sophie | Centre for Maternal, Adolescent, Reproductive and Child Health, London School of Hygiene and Tropical Medicine, London, UK |
| Sow Souleymane | Assistant de recherche, Sénégal |

